# Reliability and Validity of self-reported Vascular Risk Factors in a Multi-Ethnic Community Based Study of Aging and Dementia

**DOI:** 10.1101/2023.04.12.23288492

**Authors:** Annie J. Lee, Didi Sanchez, Dolly Reyes-Dumeyer, Adam M. Brickman, Rafael A. Lantigua, Badri N. Vardarajan, Richard Mayeux

## Abstract

**INTRODUCTION:** The reliability and validity of self-reported cardiovascular and cerebrovascular risk factors remains inconsistent in aging research.

**METHODS:** We assessed the reliability, validity, sensitivity, specificity, and percent agreement of self-reported hypertension, diabetes, and heart disease, in comparison with direct measures of blood pressure, hemoglobin A1c (HbA1c), and medication use in 1870 participants in a multiethic study of aging and dementia.

**RESULTS:** Reliability of self-reported for hypertension, diabetes, and heart disease was excellent. Agreement between self-reports and clinical measures was moderate for hypertension (kappa: 0.58), good for diabetes (kappa: 0.76-0.79), and moderate for heart disease (kappa: 0.45) differing slightly by age, sex, education, and race/ethnic group. Sensitivity and specificity for hypertension was 88.6%-78.1%, for diabetes was 87.7%-92.0% (HbA1c≥6.5%) or 92.7%-92.8% (HbA1c ≥7%), and for heart disease was 85.8%-75.5%.

**DISCUSSION:** Self-reported history of hypertension, diabetes, and heart disease are reliable and valid compared to direct measurements or medication use.

## Introduction

Cardio- and cerebrovascular risk factors are frequent among elderly adults and their presence is associated with increased risk of Alzheimer’s disease^1,2^. Self-reported questionnaires are often used to gather information about antecedent risk factors, including cardiovascular and cerebrovascular risk, for Alzheimer’s disease in observational studies^2,3^ due to convenience and lower cost relative to clinical diagnosis^4^. It is the only feasible way of obtaining information on the disease in the absence of clinical or medical records. However, the reliability, validity and overall accuracy of self-reports may be affected by the participant’s understanding of the diagnosis, willingness to report it, and ability to recall their personal information, which can be of concern among elderly individuals^4^. Well-validated and reliable self-reported information of disease risks is important for understanding the cause of the disease and may provide clues to preventive strategies.

Hypertension, diabetes, and heart disease are major cardiovascular risk factors leading increased burden of cardiovascular and cerebrovascular diseases^5,6^ and risk of dementia and Alzheimer’s disease^1,2^. The validity of self-reported information in these vascular risk factors assessed from several studies^3,4,7-15^ were found to be uncertain and highly variable reflecting differences in study population, sociodemographic characteristics, and the use of clinical measurements or treatments. There is limited information available on the reliability and validity of self-reported hypertension, diabetes, and heart disease among racially and ethnically diverse older adults.

The Washington Heights, Hamilton Heights, Inwood Columbia Aging Project (WHICAP) is a community-based longitudinal study of aging and dementia in elderly individuals living in northern Manhattan^16,17^. This cohort includes older adults who identify as non-Hispanic Black or African American, Caribbean Hispanic, and non-Hispanic White, and it also provides an opportunity assess differences by educational level, sex, clinical and genetic factors in relation to age-related diseases^18^. We previously reported higher prevalence and incidence rates of cerebrovascular disease and dementia among non-Hispanic Black and Hispanic older adults compared to non-Hispanic white individuals in WHICAP^17,19^. Thus, understanding whether the reliability and validity of self-reported vascular risk factors differs by racial and race/ethnic group is an important objective.

Among older participants in this multi-ethnic community-based WHICAP cohort, the validity of self-reported hypertension, diabetes, and heart disease was investigated using Cohen’s kappa, sensitivity, specificity, and percent agreement by comparing participant responses to direct measurements or reviewing disease-specific medications use. We also investigated the reliability of self-reports using longitudinal data, and whether the validity of self-reported hypertension, diabetes, and heart disease differed by age, sex, education, or race/ethnic group.

## Methods

### Participants

Participants were from WHICAP, a community-based longitudinal study of aging and dementia in a multiethnic cohort of individuals aged 65 years or older residing in northern Manhattan^16,17^. Participants were initially recruited as non-demented by self-report in waves in 1992, 1999, and 2009 using similar sampling strategies, assessments, and study design and procedures^17^.

### Consent Statement

The study was approved by the Institutional Review Boards of Columbia University. All participants provided written informed consent. A detailed description of the study was previously published^16^.

### Cardio- and Cerebrovascular Risk Factors (Vascular Risk Factors)

At the initial and each follow-up visit, self-reported medical history, self-reported use of disease-specific medications, and physical examinations were recorded. Self-reported information on hypertension, diabetes, and heart disease was obtained from each participant at last visit by three questions “Have you ever had a hypertension / diabetes / heart disease?” (Ever Had or Never Had).

Clinical measures were also obtained by direct measurement in a subset of the most recent WHICAP participants and from available information obtained at the interviews. For hypertension we used two approaches. First, we used the measurement of blood pressure defined as systolic blood pressure ≥140 mmHg, diastolic blood pressure ≥90 mmHg^20^, based on the WHO guidelines^21^. If no measurement of blood pressure was available, we used information regarding the use of anti-hypertension medications. For diabetes we considered two reference definitions: *definition 1:* measurement of hemoglobin A1C level ≥6.5% at last visit or the use of any medications to manage diabetes at the initial or follow-up visits; *definition 2:* hemoglobin A1C level ≥7% at last visit or diabetes medication use at the initial or follow-up visits. Cut-points for hemoglobin A1C level were based on the current American Diabetes Association guidelines^22^ (≥ 6.5%) and current Department of Veterans Affairs’ recommendations (≥7%)^23,24^. For heart disease we relied entirely on the use of medications to manage heart disease at the initial or follow-up visits. The use of medications was classified as follows: *Hypertension*: angiotensin-converting enzyme inhibitors, beta-blockers, calcium channel blockers, or diuretics; *Diabetes*: glitazones, insulin, metformin, oral hypoglycemics, or sulfonylurea; *Heart disease*: digitalis or digoxin, anti-anginal agents, nitrates, or other anti-arrhythmics/anginals. The reported use of medications was recorded as “Taken or Not Taken”.

### Covariates

Information on age at last visit, sex, education and race/ethnic group were queried. Years of education was self-reported and ranged from 0 to 20. The determination of race/ethnic group was self-reported using the 2000 US Census^25^ as a guide. We excluded the small number of individuals who did not identify as non-Hispanic Black, Caribbean Hispanic or non-Hispanic white (n = 24).

### Statistical analyses

Reliability^26,27^ of self-reported hypertension, diabetes, and heart disease was assessed at the individual level for participants with at least two visits by fitting a mixed effects logistic regression model with fixed effects including age, sex, educational level, and ethnicity and a random intercept for each individual using rpt function in R package rptR^26^. We assessed the reliability of each self-reported vascular factor beginning from the first interview at which there was an affirmative response We also investigated the reliability of self-reported risk factors at the individual level for participants with at least two visits excluding the first negative reports if they later had a positive self-report. The repeatability value ranges from 0 to 1 and the value of less than 0.40 was considered poor, 0.40-0.59 as fair, 0.60-0.74 as good, and 0.75-1.00 as excellent reliability.

Validity of self-reported risk factors data was compared to measured hypertension, diabetes and heart disease using Cohen’s kappa^28^, sensitivity^29^, specificity^29^, and percent agreement^28^. Cohen’s kappa was used to measure the agreement between self-reported and measured risk factor by taking into account the agreement expected to occur by chance. The kappa value ranges from -1 to 1 and the value of less than or equal to 0 was considered no, 0.01-0.40 as poor-to-fair, 0.41-0.60 as moderate, 0.61-0.80 as good, and 0.81-1.00 as excellent agreement^28^. Sensitivity was defined as the proportion of participants who self-reported to ever had a risk factor among those with positive measured risk factor. Specificity was defined as the proportion of participants who self-reported to never had a risk factor in whom we found no evidence of the measured risk factor. False negative rate (1-sensitivity) and false positive rate (1-specificity) were used to examine the proportion of under-reported and over-reported, respectively, for risk factors. Percent agreement was defined as the proportion of all participants with positive self-report with positive measured risk factor or negative self-report with negative measured risk factor ^28^. We compared the percent agreement for hypertension, diabetes, and heart disease.

Stratified analyses of the validity measures for self-reported hypertension, diabetes, and heart disease were subsequently compared across several demographic variables (age, sex, education, and race/ethnic group). Participants were stratified by median age of 80.9 years, younger (65 years ≤ age ˂80.9 years) and older (age ≥ 80.9 years). Participants were categorized into three groups of educational level using cut-points based on the education quartiles of the samples, low (education < 6), medium (6 ≤ education < 12), and high (education ≥ 12 years of education)^30^. Exact method^31-33^ was used to compute the confidence intervals. Kappa statistic and its confidence interval were computed using epi.kappa function in R package epiR^34^. Sensitivity and specificity and its confidence intervals were computed using BDtest function in R package bdpv^35^. Difference between two proportions was tested using two-proportions z-test using prop.test function in R package stats^36^. A p-value less than 0.05 was considered significant. The false discovery rate (FDR) of 0.05 was used to correct for multiple testing. All statistical analyses were performed using R^36^ version 4.1.3.

### Funding

All support for this work was provided through grants from the National Institue on Aging of the National Institutes of Health.

## Results

A total of 1,870 participants were 65 years or older and had complete data on self-reported and measured hypertension, diabetes and heart disease, in addition to age at last visit, sex, education, and race/ethnic group. The demographics characteristics of this cohort are in **Table 1**. The frequency of self-reported vascular risk factors: hypertension, diabetes, and heart disease are also listed in **Table 1** and **Supplementary Table 1**. The clinical measures showed a higher frequency of hypertension (p<0.001) and lower frequencies for diabetes (p=0.030 for *definition 1* and p=0.003 for *definition 2*) and heart disease (p<0.001) than the self-reported assessments.

**Table 1.** Participant demographics and clinical characteristics of the study sample.

| Characteristic | Total Sample<br>(n=1,870) |
| --- | --- |
| Age (years), mean, median (SD) | 81.16, 80.9 (7.18) |
| 65 ≤ Age < 80.9, n (%) | 847 (45%) |
| Age ≥ 80.9, n (%) | 1023 (55%) |
| Sex |  |
| Women, n (%) | 1212 (65%) |
| Men, n (%) | 658 (35%) |
| Education (years), mean (SD) | 9.39 (4.79) |
| Low: Education < 6, n (%) | 436 (23%) |
| Medium: 6 ≤ Education < 12, n (%) | 616 (33%) |
| High: Education ≥ 12, n (%) | 818 (44%) |
| Race/ethnic group |  |
| Non-Hispanic white, n (%) | 483 (26%) |
| African American, n (%) | 635 (34%) |
| Caribbean Hispanic, n (%) | 752 (40%) |
| Self-reported |  |
| Hypertension, n (%) | 1459 (78%) |
| Diabetes, n (%) | 462 (25%) |
| Heart disease, n (%) | 670 (36%) |
| Measured |  |
| Hypertension, n (%) | 1573 (84%) |
| Diabetes (*definition 1*), n (%) | 405 (22%) |
| Diabetes (*definition 2*), n (%) | 386 (21%) |
| Heart disease, n (%) | 346 (19%) |

SD: standard deviation; *definition 1*: hemoglobin A1C level ≥ 6.5% at last visit or the use of any medications to manage diabetes at initial or follow-up visits; *definition 2*: hemoglobin A1C level ≥ 7% at last visit or diabetes medication use at initial or follow-up visits.

The 1,870 participants had a total of 4,743 self-reported hypertension assessment and 4,739 self-reported diabetes and heart disease assessments at the initial and follow-up visits. Of those, when we restricted to participants with at least two visits, 1,307 participants had self-reported hypertension assessment with a total of 4,183 visits, 1,306 participants had self-reported diabetes with a total of 4,178 visits, and 1,307 participants had self-reported heart disease with a total of 4,179 visits (**Supplementary Table 2**). We excluded the first negative self-reports of participants when they later had positive self-report due to a change in health status, after restricting to participants with at least two visits, 1,240 participants had self-reported hypertension with a total of 3,860 visits, 1,269 participants had self-reported diabetes with a total of 3,985 visits, and 1,222 participants had self-reported heart disease with a total of 3,795 visits (**Supplementary Table 2**). The self-reported hypertension, diabetes, and heart disease were assessed from three visits per individual on average. The distribution of the proportion of positive self-reported risk factors among individuals is shown in **Supplementary Table 2**.

**Table 2.** Reliability of self-reported hypertension, diabetes, and heart disease among individuals, before and after excluding the first consecutive negative self-reports of participants when they later consistently had positive self-reports.

| Before exclusion | Hypertension | Diabetes | Heart disease |
| --- | --- | --- | --- |
| Unadjusted | 0.959 | 0.983 | 0.948 |
| Adjusted for sex, age, education, and ethnicity | 0.968 | 0.987 | 0.951 |
| After exclusion | Hypertension | Diabetes | Heart disease |
| Unadjusted | 0.987 | 0.995 | 0.986 |
| Adjusted for sex, age, education, and ethnicity | 0.987 | 0.995 | 0.986 |

The reliability of self-reports was excellent for hypertension (0.96 and 0.97), diabetes (0.98 and 0.99), and heart disease (0.95 and 0.95) unadjusted and adjusted for age, sex, education, and ethnicity, respectively (**Table 2**). The subsequent reliability of self-reports was excellent for hypertension (0.987), diabetes (0.995), and heart disease (0.986) when unadjusted and adjusted for age, sex, education, and ethnicity (**Table 2**).

The validity of self-reported against measured hypertension, diabetes, and heart disease is shown in **Table 3**. The agreement between self-reported and measured vascular risk factors was moderate for hypertension (kappa: 0.58, 95% confidence interval (CI): 0.53-0.63), good for diabetes (kappa: 0.76, 95% CI: 0.73-0.80 for *definition 1*; kappa: 0.79, 95% CI: 0.76-0.82 for *definition 2*), and moderate for heart disease (kappa: 0.45, 95% CI: 0.40-0.50). Our agreement for diabetes was higher than that of hypertension (*p*_*FDR*_=0.013 for *definition 1*; *p*_*FDR*_=0.004 for *definition 2*) and heart disease (*p*_*FDR*_<0.001 for *definition 1* and *2*). Sensitivity and specificity for hypertension was 88.6% (95% CI: 86.9-90.1) and 78.1% (95% CI: 73.0-82.7), that of diabetes or *definition 1* was 87.7% (95% CI: 84.0-90.7) and 92.7% (95% CI: 91.2-94.0), that of diabetes for *definition 2* was 92.0% (95% CI: 88.8-94.5) and 92.8% (95% CI: 91.4-94.1), and that of heart disease was 85.8% (95% CI: 81.7-89.3) and 75.5% (95% CI: 73.3-77.7), respectively, resulting in over-reporting and under-reporting of 21.9% and 11.4% for hypertension, 7.3% and 12.3% for diabetes for *definition 1*, 7.2% and 8.0% for diabetes for *definition 2*, and 24.5% and 14.2% for heart disease. Percent agreement for hypertension, diabetes, and heart disease were 87%, 92% for *definition 1* and 93% for *definition 2*, and 77%, respectively, and diabetes had a higher percent agreement than hypertension (*p*_*FDR*_<0.001) and heart disease (*p*_*FDR*_<0.001) for *definition 1* and *2*.

**Table 3.** Validity of self-reported against measured hypertension, diabetes and heart disease stratified by participants characteristics.

| Variable | Group | Hypertension |  |  |  |  |  |
| --- | --- | --- | --- | --- | --- | --- | --- |
|  |  | Cohen's kappa | Sensitivity (%) | Specificity (%) | Underreported | Overreported | %agree |
| All |  | 0.58 (0.53-0.63) | 88.6 (86.9-90.1) | 78.1 (73.0-82.7) | 11.4 (9.9-13.1) | 21.9 (17.3-27.0) | 87.0 |
| Age | 65-80.9 | 0.56 (0.49-0.62) | 85.6 (82.9-88.0) | 79.0 (72.2-84.7) | 14.4 (12.0-17.1) | 21.0 (15.3-27.8) | 84.3 |
|  | ≥80.9 | 0.60 (0.52-0.67) | 91.5 (89.4-93.3) * | 76.9 (68.3-84.0) | 8.5 (6.7-10.6) * | 23.1 (16-31.7) | 89.6 * |
| Sex | Women | 0.59 (0.52-0.65) | 90.3 (88.3-92.0) | 78.9 (71.8-84.9) | 9.7 (8.0-11.7) | 21.1 (15.1-28.2) | 88.8 |
|  | Men | 0.56 (0.48-0.63) | 85.2 (81.9-88.2) * | 77.2 (69.2-84.0) | 14.8 (11.8-18.1) * | 22.8 (16.0-30.8) | 83.6 * |
| Education | <6 | 0.51 (0.40-0.63) | 89.8 (86.2-92.7) | 67.2 (54.3-78.4) | 10.2 (7.3-13.8) | 32.8 (21.6-45.7) | 86.5 |
|  | 6-12 | 0.57 (0.48-0.66) | 89.8 (86.9-92.3) | 76.7 (66.4-85.2) | 10.2 (7.7-13.1) | 23.3 (14.8-33.6) | 88.0 |
|  | ≥12 | 0.61 (0.54-0.67) | 87.0 (84.3-89.5) | 83.7 (76.7-89.3) | 13.0 (10.5-15.7) | 16.3 (10.7-23.3) | 86.4 |
| Ethnicity | Whites | 0.63 (0.55-0.71) | 85.0 (81.0-88.4) | 89.7 (81.9-94.9) | 15.0 (11.6-19.0) | 10.3 (5.1-18.1) | 85.9 |
|  | AfAm | 0.57 (0.47-0.66) | 90.9 (88.2-93.2) | 73.3 (62.6-82.2) | 9.1 (6.8-11.8) | 26.7 (17.8-37.4) | 88.5 |
|  | Hispanics | 0.53 (0.45-0.62) | 88.9 (86.2-91.2) | 71.9 (62.7-79.9)* | 11.1 (8.8-13.8) | 28.1 (20.1-37.3)* | 86.3 |
| Variable | Group | Diabetes (*definition 1*) |  |  |  |  |  |
|  |  | Cohen's kappa | Sensitivity (%) | Specificity (%) | Underreported | Overreported | %agree |
| All |  | 0.76 (0.73-0.80) | 87.7 (84.0-90.7) | 92.7 (91.2-94.0) | 12.3 (9.3-16.0) | 7.3 (6.0-8.8) | 91.6 |
| Age | 65-80.9 | 0.78 (0.73-0.82) | 91.2 (86.6-94.6) | 91.7 (89.4-93.6) | 8.8 (5.4-13.4) | 8.3 (6.4-10.6) | 91.6 |
|  | ≥80.9 | 0.75 (0.70-0.80) | 83.6 (77.5-88.6) | 93.7 (91.7-95.3) | 16.4 (11.4-22.5) | 6.3 (4.7-8.3) | 91.6 |
| Sex | Women | 0.77 (0.72-0.81) | 86.6 (81.9-90.5) | 93.2 (91.4-94.7) | 13.4 (9.5-18.1) | 6.8 (5.3-8.6) | 91.7 |
|  | Men | 0.76 (0.70-0.82) | 89.5 (83.3-94.0) | 91.8 (89.1-94.1) | 10.5 (6.0-16.7) | 8.2 (5.9-10.9) | 91.3 |
| Education | <6 | 0.78 (0.71-0.84) | 91.6 (85.1-95.9) | 90.2 (86.4-93.3) | 8.4 (4.1-14.9) | 9.8 (6.7-13.6) | 90.6 |
|  | 6-12 | 0.75 (0.69-0.81) | 89.4 (83.1-93.9) | 90.7 (87.8-93.2) | 10.6 (6.1-16.9) | 9.3 (6.8-12.2) | 90.4 |
|  | ≥12 | 0.77 (0.71-0.82) | 82.8 (75.6-88.5) | 95.2 (93.4-96.7) * | 17.2 (11.5-24.4) | 4.8 (3.3-6.6) | 93.0 |
| Ethnicity | Whites | 0.74 (0.65-0.83) | 80.3 (68.7-89.1) | 95.7 (93.3-97.4) | 19.7 (10.9-31.3) | 4.3 (2.6-6.7) | 93.6 |
|  | AfAm | 0.74 (0.67-0.80) | 84.3 (77.2-89.9) | 92.3 (89.6-94.5) | 15.7 (10.1-22.8) | 7.7 (5.5-10.4) | 90.6 |
|  | Hispanics | 0.79 (0.74-0.84) | 92.5 (87.9-95.7) | 90.8 (88.1-93.1) * | 7.5 (4.3-12.1) | 9.2 (6.9-11.9) | 91.2 |

| Variable | Group | Diabetes (*definition 2*) |  |  |  |  |  |
| --- | --- | --- | --- | --- | --- | --- | --- |
|  |  | Cohen's kappa | Sensitivity (%) | Specificity (%) | Underreported | Overreported | %agree |
| All |  | 0.79 (0.76-0.82) | 92.0 (88.8-94.5) | 92.8 (91.4-94.1) | 8.0 (5.5-11.2) | 7.2 (5.9-8.6) | 92.6 |
| Age | 65-80.9 | 0.80 (0.75-0.84) | 94.7 (90.7-97.3) | 91.8 (89.5-93.7) | 5.3 (2.7-9.3) | 8.2 (6.3-10.5) | 92.4 |
|  | ≥80.9 | 0.78 (0.73-0.83) | 88.8 (83.2-93.0) | 93.8 (91.8-95.4) | 11.2 (7.0-16.8) | 6.2 (4.6-8.2) | 92.8 |
| Sex | Women | 0.80 (0.76-0.84) | 91.9 (87.8-95.0) | 93.3 (91.5-94.8) | 8.1 (5.0-12.2) | 6.7 (5.2-8.5) | 93.0 |
|  | Men | 0.78 (0.72-0.83) | 92.1 (86.3-96.0) | 91.9 (89.2-94.1) | 7.9 (4.0-13.7) | 8.1 (5.9-10.8) | 91.9 |
| Education | <6 | 0.80 (0.74-0.86) | 95.6 (90.1-98.6) | 90.4 (86.6-93.4) | 4.4 (1.4-9.9) | 9.6 (6.6-13.4) | 91.7 |
|  | 6-12 | 0.76 (0.70-0.82) | 91.3 (85.3-95.4) | 90.8 (87.8-93.2) | 8.7 (4.6-14.7) | 9.2 (6.8-12.2) | 90.9 |
|  | ≥12 | 0.81 (0.75-0.86) | 89.6 (83.1-94.2) | 95.3 (93.5-96.8) * | 10.4 (5.8-16.9) | 4.7 (3.2-6.5) | 94.4 * |
| Ethnicity | Whites | 0.79 (0.71-0.88) | 91.4 (81.0-97.1) | 95.8 (93.4-97.5) | 8.6 (2.9-19.0) | 4.2 (2.5-6.6) | 95.2 |
|  | AfAm | 0.76 (0.70-0.82) | 88.7 (82.1-93.5) | 92.4 (89.8-94.6) | 11.3 (6.5-17.9) | 7.6 (5.4-10.2) | 91.7 * |
|  | Hispanics | 0.80 (0.75-0.85) | 94.4 (90.1-97.2) | 90.8 (88.1-93.1) | 5.6 (2.8-9.9) | 9.2 (6.9-11.9) | 91.8 * |
| Variable | Group | Heart disease |  |  |  |  |  |
|  |  | Cohen's kappa | Sensitivity (%) | Specificity (%) | Underreported | Overreported | %agree |
| All |  | 0.45 (0.40-0.50) | 85.8 (81.7-89.3) | 75.5 (73.3-77.7) | 14.2 (10.7-18.3) | 24.5 (22.3-26.7) | 77.4 |
| Age | 65-80.9 | 0.43 (0.36-0.51) | 86.9 (79.9-92.2) | 78.5 (75.5-81.3) | 13.1 (7.8-20.1) | 21.5 (18.7-24.5) | 79.6 |
|  | ≥80.9 | 0.45 (0.39-0.51) | 85.2 (79.7-89.6) | 72.2 (68.8-75.5) * | 14.8 (10.4-20.3) | 27.8 (24.5-31.2) * | 75.2 * |
| Sex | Women | 0.45 (0.39-0.50) | 83.3 (77.8-87.9) | 76.1 (73.4-78.8) | 16.7 (12.1-22.2) | 23.9 (21.2-26.6) | 77.5 |
|  | Men | 0.46 (0.38-0.54) | 90.8 (84.1-95.3) | 74.4 (70.5-78.0) | 9.2 (4.7-15.9) | 25.6 (22.0-29.5) | 77.4 |
| Education | <6 | 0.48 (0.38-0.58) | 80.2 (70.2-88.0) | 79.4 (74.8-83.5) | 19.8 (12-29.8) | 20.6 (16.5-25.2) | 79.6 |
|  | 6-12 | 0.45 (0.37-0.53) | 87.9 (80.6-93.2) | 74.4 (70.3-78.2) | 12.1 (6.8-19.4) | 25.6 (21.8-29.7) | 76.9 |
|  | ≥12 | 0.43 (0.36-0.50) | 87.5 (81.0-92.4) | 74.3 (70.9-77.6) | 12.5 (7.6-19.0) | 25.7 (22.4-29.1) | 76.7 |
| Ethnicity | Whites | 0.42 (0.33-0.51) | 91.2 (83.4-96.1) | 70.2 (65.4-74.6) | 8.8 (3.9-16.6) | 29.8 (25.4-34.6) | 74.1 |
|  | AfAm | 0.44 (0.36-0.52) | 84.2 (75.6-90.7) | 77.9 (74.1-81.4) | 15.8 (9.3-24.4) | 22.1 (18.6-25.9) | 78.9 |
|  | Hispanics | 0.48 (0.41-0.55) | 83.8 (77.0-89.2) | 76.9 (73.3-80.2) | 16.2 (10.8-23.0) | 23.1 (19.8-26.7) | 78.3 |

*Whites*: Non-Hispanic Whites; *AfAm*: African Americans; %agree: Percent agreement; *definition 1*: hemoglobin A1C level ≥6.5% at last visit or the use of any medications to manage diabetes at initial or follow-up visits; *definition 2*: hemoglobin A1C level ≥7% at last visit or diabetes medication use at initial or follow-up visits. \* significant at p-value <0.05 or $p_{FDR}$ <0.05 for a comparison within a group of 3 strata.

When stratified by sex, age, education, or race/ethnic group, the sensitivity of hypertension was higher for older, than younger adults (91.5% vs. 85.6%, p=0.0003) and higher for women, than men (90.3% vs. 85.2%, p=0.004) (**Table 3**). Specificity of hypertension was higher for non-Hispanic Whites, than Hispanics (89.7% vs. 71.9%, *p*_*FDR*_=0.007), that of diabetes was higher for participants with high education, than low education (95.2% vs. 90.2%, *p*_*FDR*_=0.006 for *definition 1*; 95.3% vs. 90.4%, *p*_*FDR*_=0.006 for *definition 2*), that of diabetes was higher for non-Hispanic white individuals, than Hispanics (95.7% vs. 90.8%, *p*_*FDR*_=0.015 for *definition 1*; 95.8% vs. 90.8%, *p*_*FDR*_=0.013 for *definition 2*), and that of heart disease was lower for older, than younger adults (72.2% vs. 78.5%, p=0.006). Percent agreement of hypertension was higher for older, than younger adults (89.6% vs. 84.3%, p=0.001) and higher for women, than men (88.8% vs. 83.6, p=0.002), that of diabetes was higher for participants with high education, than medium education (94.4% vs. 90.9%, *p*_*FDR*_=0.046 for *definition 2*) and higher for non-Hispanic white individuals, than non-Hispanic Black participants and Hispanics (95.2% vs. 91.7%, *p*_*FDR*_=0.038; 95.2% vs. 91.8%, *p*_*FDR*_=0.038 for *definition* 2), that of heart disease was lower for older, than younger adults (75.2% vs. 79.6%; p=0.026). There were no significant differences in Cohen’s kappa, sensitivity, specificity, and percent agreement of the risk factors for the other strata.

## Discussion

This investigation assessed the reliability of self-reported vascular risk factors and the validity when compared with directly measured hypertension, diabetes, and heart disease in a community-based study of older individuals of African and Hispanic ancestry, and non-Hispanic white individuals of European ancestry. We found excellent reliability of self-reported hypertension, diabetes, and heart disease with and without adjustment for age, sex, education, and ethnicity. We also found good agreement between self-reports of vascular risk factors and measured HbA1c, and moderate agreement for measured hypertension and medication use for heart disease. Sensitivity, specificity, and percent agreement were high for diabetes and moderate for hypertension and heart disease. In stratified analyses, sensitivity and percent agreement of hypertension were highest in older adults and women. The specificity of hypertension and diabetes was slightly higher in non-Hispanic white individuals compared with Hispanic individuals. Individuals with more education also had higher specificity for diabetes. The specificity of heart disease was lowest for the oldest individuals. Percent agreement of diabetes for was higher among individuals with more education, and higher in non-Hispanic white individuals, than African-Americans or Hispanics.

The agreement for hypertension was consistent with prior studies in older adults^8,10-12^, as was the sensitivity^7,9,11,13^. The sensitivity for non-Hispanic white (85%) and non-Hispanic black individuals (91%), and Hispanics (89%) were similar to previously reported older US-born adults^13^. The specificity and the percent agreement of self-reported hypertension was consistent with prior studies^7,9,11,13^. It is possible that these participants were 15-20 years older than in previous studies^7,9,13^ and perhaps more accustomed to reporting the presence of hypertension^37-39^.

For diabetes, reliability as measured by the kappa for both categories of HbA1c was consistent with prior studies in older adults^8,11,12,40^. The sensitivity, specificity percent agreement were similar to prior studies^4,9,11,12,40,41^, but only a few studies compared self-reported diabetes with hemoglobin HbA1c^8,12,40^. The intensive intervention, lifestyle management in terms of diet and exercise, and use of medications required for diabetes may explain the higher awareness of diabetes^11^.

The agreement for heart disease was consistent with previous studies^10-12^ of myocardial infarction and heart failure^7,9-12^. Self-reported heart failure has been difficult to validate compared with clinical measures throughout the literature^42^. The low agreement may reflect the complexity of diagnosing and classifying various types of heart disease and conveying these diagnosis to patients^42^. Despite these limitations, the sensitivity was reasonable and within the range of previous reports on myocardial infarction and heart failure^7,9,11,12^. The specificity and the agreement were lower than that observed in previous studies^7,9,11,12^. A possible explanation for the difference was the definition of heart disease. Medical documentation was not available for the heart disease in this study. This required that we rely entirely on the reported use of medications to manage all types of heart disease. Another explanation could be limited knowledge about types of heart disease^12^. It has been shown that awareness for heart failure is lower than for other heart conditions in the general population^43^.

Previous studies have investigated the validity of self-reported cardiovascular risk factors in populations at risk for stroke^4,18^. Similar to our investigation, data were extracted from questionnaires and compared with direct measurements or medical records. Overall accuracy was best for hypertension and diabetes, but not for hypercholesterolemia. The sensitivities for hypertension reported in other studies reviewed by Bowlin et al^44^ were slightly lower than those reported here. However, high sensitivity and specificity were found in older non-Hispanic Blacks, Hispanics and non-Hispanic Whites in the Health and Retirement Study^13^. This suggests that the accuracy of self-reported vascular risk factors is variable but not limiting.

Strengths of our study include the use of a large, well-characterized, longitudinal, multi-ethnic cohort that allowed us to evaluate the reliability and validity of self-reported risk factors among a diverse group of older adults. Moreover, the use of HbA1c to assess the validity of self-reported diabetes is important because using this measure is recommended for the diagnosis of diabetes by the American Diabetes Association^22^.

Despite these strengths, our study is subject to potential limitations. The study was conducted in older population aged 65 years or older living in northern Manhattan with a high frequency of cardio- and cerebrovascular risk factors. This may have limited generalizability to other age groups or cohorts with lower morbidity. The medical documentation of heart disease was not available and we relied entirely on self-reported medication use. This may have resulted in lower specificity and lower percent agreement than the previous literatures. Hispanic individuals have fewer years of education than non-Hispanic Black and non-Hispanic White individuals, which can affect the accuracy of self-report^45^.

Our results indicate that there is excellent reliability among older individuals for self-reported hypertension, diabetes, and heart disease. Furthermore, agreement, sensitivity, specificity, and percent agreement of self-reported diabetes were good and that of hypertension and heart disease were moderate when using clinical measures as validation. Establishing reliability and validity will also augment efforts to harmonize data across similar epidemiological studies.

## Supporting information

CVRF.supplement files.040923

## Data Availability

All data produced in the present study are available upon reasonable request to the authors
All data produced in the present work are contained in the manuscript

## Acknowledgements

This research is supported by National Institutes of Health grants R01AG037212, R01AG072474, R25GM143298, and U24AG074855.

## Contributors

A.J.L. and R.M. created the study concept and designed the study. A.J.L., B.N.V., and R.M. interpreted the experiments. A.J.L., B.N.V., and R.M. drafted the manuscript. A.J.L. performed and analyzed the experiments. All authors contributed to data acquisition and interpretation and critically reviewed the manuscript. All authors have read and approved the manuscript.

## RESEARCH IN CONTEXT

### Systematic Review

The authors reviewed the epidemiological literature on the use of questionnaires for cardiovascular and cerebrovascular risk factors. These vascular risk factors are important antecedents for heart disease, stroke, and dementia. However, the reliability and validity of self-reported vascular risk factors has not been widely investigated in studies of aging and dementia.

### Interpretation

The findings here show that self-report of the most common vascular risk factors is reliable and valid. However, there are slight, but important differences by age, sex, education, and race/ethnic group in validity of these self-reported conditions when compared with direct measurements.

### Future directions

This study provides confidence that ascertainment of vascular risk factors through self-report is reliable and valid. Establishing reliable and valid questionnaires across different race/ethnic groups, languages, educational levels, age, and sex is also critically important in the study of dementia.

Click here to access/download

Supplementary Files

Supplemental Data.CVRF.032723.docx

## References

1. Luchsinger JA, Mayeux R. Cardiovascular risk factors and Alzheimer’s disease. Curr Atheroscler Rep. 2004;6(4):261–266.

2. Tosto G, Bird TD, Bennett DA, et al. The Role of Cardiovascular Risk Factors and Stroke in Familial Alzheimer Disease. JAMA Neurol. 2016;73(10):1231–1237.

3. Haapanen N, Miilunpalo S, Pasanen M, Oja P, Vuori I. Agreement between questionnaire data and medical records of chronic diseases in middle-aged and elderly Finnish men and women. Am J Epidemiol. 1997;145(8):762–769.

4. Dey AK, Alyass A, Muir RT, et al. Validity of Self-Report of Cardiovascular Risk Factors in a Population at High Risk for Stroke. J Stroke Cerebrovasc Dis. 2015;24(12):2860–2865.

5. Yusuf S, Reddy S, Ounpuu S, Anand S. Global burden of cardiovascular diseases: part I: general considerations, the epidemiologic transition, risk factors, and impact of urbanization. Circulation. 2001;104(22):2746–2753.

6. Vaduganathan M, Mensah GA, Turco JV, Fuster V, Roth GA. The Global Burden of Cardiovascular Diseases and Risk: A Compass for Future Health. J Am Coll Cardiol. 2022;80(25):2361–2371.

7. Okura Y, Urban LH, Mahoney DW, Jacobsen SJ, Rodeheffer RJ. Agreement between self-report questionnaires and medical record data was substantial for diabetes, hypertension, myocardial infarction and stroke but not for heart failure. J Clin Epidemiol. 2004;57(10):1096–1103.

8. Hansen H, Schafer I, Schon G, et al. Agreement between self-reported and general practitioner-reported chronic conditions among multimorbid patients in primary care - results of the MultiCare Cohort Study. BMC Fam Pract. 2014;15:39.

9. Englert H, Muller-Nordhorn J, Seewald S, et al. Is patient self-report an adequate tool for monitoring cardiovascular conditions in patients with hypercholesterolemia? J Public Health (Oxf). 2010;32(3):387–394.

10. Teh R, Doughty R, Connolly M, et al. Agreement between self-reports and medical records of cardiovascular disease in octogenarians. J Clin Epidemiol. 2013;66(10):1135–1143.

11. Steinkirchner AB, Zimmermann ME, Donhauser FJ, et al. Self-report of chronic diseases in old-aged individuals: extent of agreement with general practitioner medical records in the German AugUR study. J Epidemiol Community Health. 2022;76(11):931–938.

12. Ryden L, Sigstrom R, Nilsson J, et al. Agreement between self-reports, proxy-reports and the National Patient Register regarding diagnoses of cardiovascular disorders and diabetes mellitus in a population-based sample of 80-year-olds. Age Ageing. 2019;48(4):513–518.

13. White K, Avendano M, Capistrant BD, Robin Moon J, Liu SY, Maria Glymour M. Self-reported and measured hypertension among older US- and foreign-born adults. J Immigr Minor Health. 2012;14(4):721–726.

14. Klungel OH, de Boer A, Paes AH, Seidell JC, Bakker A. Cardiovascular diseases and risk factors in a population-based study in The Netherlands: agreement between questionnaire information and medical records. Neth J Med. 1999;55(4):177–183.

15. Alonso A, Beunza JJ, Delgado-Rodriguez M, Martinez-Gonzalez MA. Validation of self reported diagnosis of hypertension in a cohort of university graduates in Spain. BMC Public Health. 2005;5:94.

16. Tang MX, Stern Y, Marder K, et al. The APOE-epsilon4 allele and the risk of Alzheimer disease among African Americans, whites, and Hispanics. JAMA. 1998;279(10):751–755.

17. Tang MX, Cross P, Andrews H, et al. Incidence of AD in African-Americans, Caribbean Hispanics, and Caucasians in northern Manhattan. Neurology. 2001;56(1):49–56.

18. Reitz C, Schupf N, Luchsinger JA, et al. Validity of self-reported stroke in elderly African Americans, Caribbean Hispanics, and Whites. Arch Neurol. 2009;66(7):834–840.

19. Brickman AM, Schupf N, Manly JJ, et al. Brain morphology in older African Americans, Caribbean Hispanics, and whites from northern Manhattan. Arch Neurol. 2008;65(8):1053–1061.

20. Chobanian AV, Bakris GL, Black HR, et al. Seventh report of the Joint National Committee on Prevention, Detection, Evaluation, and Treatment of High Blood Pressure. Hypertension. 2003;42(6):1206–1252.

21. Whelton PK, Carey RM, Aronow WS, et al. 2017 ACC/AHA/AAPA/ABC/ACPM/AGS/APhA/ASH/ASPC/NMA/PCNA Guideline for the Prevention, Detection, Evaluation, and Management of High Blood Pressure in Adults: Executive Summary: A Report of the American College of Cardiology/American Heart Association Task Force on Clinical Practice Guidelines. Hypertension. 2018;71(6):1269–1324.

22. American Diabetes A. Diagnosis and classification of diabetes mellitus. Diabetes Care. 2010;33 Suppl 1(Suppl 1):S62–69.

23. Tschanz MP, Watts SA, Colburn JA, Conlin PR, Pogach LM. Overview and Discussion of the 2017 VA/DoD Clinical Practice Guideline for the Management of Type 2 Diabetes Mellitus in Primary Care. Fed Pract. 2017;34(Suppl 8):S14–S19.

24. U.S. Department of Veteran Affairs, U.S. Department of Defense. VA/DoD clinical practice guidelines: management of diabetes mellitus in primary care. [Accessed August 28, 2017].https://www.healthquality.va.gov/guidelines/CD/diabetes/. Updated April 18, 2017.

25. Census UBOT. Census of Population and Housing Summary Tape File1, Technical Documentation. WAshington, DC. 1990.

26. Stoffel MA, Nakagawa S, Schielzeth H. rptR: repeatability estimation and variance decomposition by generalized linear mixed-effects models. Methods Ecol Evol. 2017;8(11):1639–1644.

27. Chervet N, Zottl M, Schurch R, Taborsky M, Heg D. Repeatability and heritability of behavioural types in a social cichlid. Int J Evol Biol. 2011;2011:321729.

28. McHugh ML. Interrater reliability: the kappa statistic. Biochem Med (Zagreb). 2012;22(3):276–282.

29. Parikh R, Mathai A, Parikh S, Chandra Sekhar G, Thomas R. Understanding and using sensitivity, specificity and predictive values. Indian J Ophthalmol. 2008;56(1):45–50.

30. Bertola L, Wei-Ming Watson C, Avila JF, et al. Predictors of Episodic Memory Performance Across Educational Strata: Multiple-Group Comparisons. J Int Neuropsychol Soc. 2019;25(9):901–909.

31. Clopper CJ, Pearson ES. The use of confidence or fiducial limits illustrated in the case of the binomial. Biometrika. 1934;26(4):404–413.

32. Searle SR. Linear Models. Vol 16. Biometrische Zeitschrift: John Wiley & Sons, Ltd; 2007.

33. Collett D. Modelling Binary Data. New York Chapman and Hall/CRC; 1999.

34. Stevenson M, Nunes T, Heuer C, et al. epiR: Tools for the Analysis of Epidemiological Data. 2018;R package version 2.0.52.

35. Schaarschmidt F. bdpv: Inference and Design for Predictive Values in Diagnostic Tests. 2019;R package version 1.3.

36. R Core Team. R: A Language and Environment for Statistical Computing. R Foundation for Statistical Computing. 2022.

37. Thomas SJ, Booth JN, 3rd, Dai C, et al. Cumulative Incidence of Hypertension by 55 Years of Age in Blacks and Whites: The CARDIA Study. J Am Heart Assoc. 2018;7(14).

38. Fuchs FD. Why do black Americans have higher prevalence of hypertension?: an enigma still unsolved. Hypertension. 2011;57(3):379–380.

39. Lackland DT. Racial differences in hypertension: implications for high blood pressure management. Am J Med Sci. 2014;348(2):135–138.

40. Schneider AL, Pankow JS, Heiss G, Selvin E. Validity and reliability of self-reported diabetes in the Atherosclerosis Risk in Communities Study. Am J Epidemiol. 2012;176(8):738–743.

41. Ning M, Zhang Q, Yang M. Comparison of self-reported and biomedical data on hypertension and diabetes: findings from the China Health and Retirement Longitudinal Study (CHARLS). BMJ Open. 2016;6(1):e009836.

42. Camplain R, Kucharska-Newton A, Loehr L, et al. Accuracy of Self-Reported Heart Failure. The Atherosclerosis Risk in Communities (ARIC) Study. J Card Fail. 2017;23(11):802–808.

43. Remme WJ, McMurray JJ, Rauch B, et al. Public awareness of heart failure in Europe: first results from SHAPE. Eur Heart J. 2005;26(22):2413–2421.

44. Bowlin SJ, Morrill BD, Nafziger AN, Lewis C, Pearson TA. Reliability and changes in validity of self-reported cardiovascular disease risk factors using dual response: the behavioral risk factor survey. J Clin Epidemiol. 1996;49(5):511–517.

45. Zajacova A, Lawrence EM. The Relationship Between Education and Health: Reducing Disparities Through a Contextual Approach. Annu Rev Public Health. 2018;39:273–289.

