## Supplementary material for "Reliability and Validity of self-reported Vascular Risk Factors in a Multi-Ethnic Community Based Study of Aging and Dementia": CVRF.supplement files.040923: CVRF.supplement files.040923.docx

**Supplemental Table 1.** Frequencies of self-reported and measured hypertension, diabetes, and heart disease.

| **Hypertension** |  | Measured | | |
| --- | --- | --- | --- | --- |
|  |  | Yes | No | Total |
| Self-reported | Yes | 1394 (89%) | 65 (22%) | 1459 (78%) |
|  | No | 179 (11%) | 232 (78%) | 411 (22%) |
|  | Total | 1573 (100%) | 297 (100%) | 1870 (100%) |
| **Diabetes (*definition 1*)** |  | Measured | | |
|  |  | Yes | No | Total |
| Self-reported Diabetes | Yes | 355 (88%) | 107 (7%) | 462 (25%) |
|  | No | 50 (12%) | 1358 (93%) | 1408 (75%) |
|  | Total | 405 (100%) | 1465 (100%) | 1870 (100%) |
| **Diabetes (*definition 2*)** |  | Measured | | |
|  |  | Yes | No | Total |
| Self-reported | Yes | 355 (92%) | 107 (7%) | 462 (25%) |
|  | No | 31 (8%) | 1377 (93%) | 1408 (75%) |
|  | Total | 386 (100%) | 1484 (100%) | 1870 (100%) |
| **Heart disease** |  | Measured | | |
|  |  | Yes | No | Total |
| Self-reported | Yes | 297 (86%) | 373 (19%) | 670 (36%) |
|  | No | 49 (14%) | 1151 (81%) | 1200 (64%) |
|  | Total | 346 (100%) | 1524 (100%) | 1870 (100%) |

*definition 1*: hemoglobin A1C level $\geq$6.5% at last visit or the use of any medications to manage diabetes at initial or follow-up visits; *definition 2*: hemoglobin A1C level $\geq$7% at last visit or diabetes medication use at initial or follow-up visits.

**Supplemental Table 2.** Distribution of the proportion of positive self-reported hypertension, diabetes, and heart disease among individuals in longitudinal WHICAP cohort, before and after excluding the first consecutive negative self-reports of participants when they later consistently had positive self-reports.

|  | Before exclusion | | | | | | After exclusion | | | | | |
| --- | --- | --- | --- | --- | --- | --- | --- | --- | --- | --- | --- | --- |
|  | Hypertension | | Diabetes | | Heart Disease | | Hypertension | | Diabetes | | Heart Disease | |
| Mean | n | % | n | % | n | % | n | % | n | % | n | % |
| 0.00 | 232 | 17.75 | 962 | 73.66 | 779 | 59.60 | 232 | 18.71 | 962 | 75.81 | 779 | 63.75 |
| 0.14 | 1 | 0.08 | 1 | 0.08 | 1 | 0.08 | 0 | 0.00 | 0 | 0.00 | 1 | 0.08 |
| 0.17 | 3 | 0.23 | 6 | 0.46 | 3 | 0.23 | 1 | 0.08 | 1 | 0.08 | 0 | 0.00 |
| 0.20 | 7 | 0.54 | 5 | 0.38 | 11 | 0.84 | 1 | 0.08 | 2 | 0.16 | 1 | 0.08 |
| 0.22 | 0 | 0.00 | 0 | 0.00 | 1 | 0.08 | 0 | 0.00 | 0 | 0.00 | 1 | 0.08 |
| 0.25 | 1 | 0.08 | 5 | 0.38 | 16 | 1.22 | 0 | 0.00 | 3 | 0.24 | 4 | 0.33 |
| 0.29 | 3 | 0.23 | 1 | 0.08 | 8 | 0.61 | 1 | 0.08 | 0 | 0.00 | 5 | 0.41 |
| 0.30 | 1 | 0.08 | 0 | 0.00 | 0 | 0.00 | 1 | 0.08 | 0 | 0.00 | 0 | 0.00 |
| 0.33 | 28 | 2.14 | 18 | 1.38 | 33 | 2.52 | 4 | 0.32 | 9 | 0.71 | 7 | 0.57 |
| 0.40 | 7 | 0.54 | 4 | 0.31 | 15 | 1.15 | 3 | 0.24 | 1 | 0.08 | 14 | 1.15 |
| 0.43 | 2 | 0.15 | 4 | 0.31 | 2 | 0.15 | 1 | 0.08 | 2 | 0.16 | 2 | 0.16 |
| 0.50 | 53 | 4.06 | 25 | 1.91 | 70 | 5.36 | 12 | 0.97 | 1 | 0.08 | 15 | 1.23 |
| 0.57 | 6 | 0.46 | 0 | 0.00 | 3 | 0.23 | 2 | 0.16 | 0 | 0.00 | 2 | 0.16 |
| 0.60 | 5 | 0.38 | 5 | 0.38 | 9 | 0.69 | 3 | 0.24 | 4 | 0.32 | 6 | 0.49 |
| 0.62 | 1 | 0.08 | 0 | 0.00 | 0 | 0.00 | 1 | 0.08 | 0 | 0.00 | 0 | 0.00 |
| 0.67 | 63 | 4.82 | 26 | 1.99 | 84 | 6.43 | 20 | 1.61 | 7 | 0.55 | 51 | 4.17 |
| 0.70 | 1 | 0.08 | 0 | 0.00 | 0 | 0.00 | 1 | 0.08 | 0 | 0.00 | 0 | 0.00 |
| 0.71 | 2 | 0.15 | 1 | 0.08 | 3 | 0.23 | 2 | 0.16 | 1 | 0.08 | 2 | 0.16 |
| 0.75 | 25 | 1.91 | 5 | 0.38 | 18 | 1.38 | 9 | 0.73 | 1 | 0.08 | 10 | 0.82 |
| 0.78 | 1 | 0.08 | 0 | 0.00 | 0 | 0.00 | 1 | 0.08 | 0 | 0.00 | 0 | 0.00 |
| 0.80 | 13 | 0.99 | 4 | 0.31 | 6 | 0.46 | 10 | 0.81 | 1 | 0.08 | 5 | 0.41 |
| 0.83 | 10 | 0.77 | 3 | 0.23 | 1 | 0.08 | 8 | 0.65 | 2 | 0.16 | 1 | 0.08 |
| 0.86 | 6 | 0.46 | 2 | 0.15 | 3 | 0.23 | 2 | 0.16 | 1 | 0.08 | 2 | 0.16 |
| 0.88 | 1 | 0.08 | 0 | 0.00 | 0 | 0.00 | 1 | 0.08 | 0 | 0.00 | 0 | 0.00 |
| 0.89 | 1 | 0.08 | 0 | 0.00 | 0 | 0.00 | 1 | 0.08 | 0 | 0.00 | 0 | 0.00 |
| 0.90 | 1 | 0.08 | 0 | 0.00 | 0 | 0.00 | 0 | 0.00 | 0 | 0.00 | 0 | 0.00 |
| 1.00 | 833 | 63.73 | 229 | 17.53 | 241 | 18.44 | 923 | 74.44 | 271 | 21.36 | 314 | 25.70 |
| Total | 1307 | 100 | 1306 | 100 | 1307 | 100 | 1240 | 100 | 1269 | 100 | 1222 | 100 |

*Mean*: number of positive self-reported risk factor over the number of visits within an individual; *n*: number of individuals; *%*: percentage of individuals.
